# Dicere Non Nocere: Public Disclosure of Identifiable Patient Information by Health Professionals on Social Media

**DOI:** 10.1101/2020.03.05.20031526

**Authors:** Matthew S. Katz, Wasim Ahmed, Thomas G. Gutheil, Reshma Jagsi

**Affiliations:** Department of Radiation Medicine, Lowell General Hospital, Lowell, MA; Newcastle University Business School, Newcastle University, Newcastle Upon Tyne, United Kingdom; Department of Psychiatry, Beth Israel Deaconess Medical Center, Boston MA; Massachusetts Mental Health Center, Harvard Medical School, Boston, MA; Center for Bioethics and Social Science in Medicine, University of Michigan, Ann Arbor, MI

## Abstract

**Background:** Respecting patient privacy and confidentiality is critical for doctor-patient relationships and public trust in medical professionals. The frequency of potentially identifiable disclosures online during periods of active engagement is unknown. Our aim was to quantify potentially identifiable content shared by physicians and other health care providers on social media using the hashtag #ShareAStoryInOneTweet.

**Methods:** We used Symplur Signals software to access Twitter’s API and searched for tweets including the hashtag. We identified 1206 tweets by doctors, nurses, and other health professionals out of 43,374 tweets shared May 1-31, 2018. We evaluated tweet content in January 2019, eight months after the study period. To determine the incidence of sharing names or potentially identifiable information about patients, we performed a content analysis of the 754 tweets in which tweets disclosed information about others. We also evaluated whether participants raised concerns about privacy breaches and estimated the frequency of deleted tweets. We used dual, blinded coding for a 10% sample to estimate inter-coder reliability for potential identifiability of tweet content using Cohen’s kappa statistic.

**Results:** 656 participants, including 486 doctors (74.1%) and 98 nurses (14.9%), shared 754 tweets disclosing information about others rather than themselves. Professional participants sharing stories about patient care disclosed the time frame in 95 (12.6%) and included patient names in 15 (2.0%) of tweets. We estimated that friends or families could likely identify the clinical scenario described in 32.1% of the 754 tweets. Among 348 tweets about potentially living patients, we estimated 162 (46.6%) were likely identifiable by patients. Inter-coder reliability in rating the potential identifiability demonstrated 86.8% agreement, with a Cohen’s Kappa of 0.8 suggesting substantial agreement Of the 1206 tweets we identified, 78 (6.5%) had been deleted on the website but were still viewable in the analytics software dataset.

**Conclusions:** During periods of active sharing online, nurses, physicians, and other health professionals may sometimes share more information than patients or families might expect. More study is needed to determine whether similar events arise frequently online and to understand how to best ensure that patients’ rights are adequately respected.

## Introduction

Physicians, nurses, and other health professionals remain among the most trusted professions in the U.S. because of their commitment to the well being of others, making them a source of trusted health information and guidance.^1^ Surveys demonstrate the high trust of the U.S. public in health care professionals (HCPs) with even higher levels of trust in other countries.^1,-3^ Still recited by many medical students as they become physicians, the Hippocratic oath reflects the fundamental importance of patient privacy as a critical element of the doctor-patient relationship and precondition for the trust of the public. In the U.S., the Health Insurance Portability and Accountability Act (HIPAA) requires de-identification to avoid sharing protected health information.^4^

Fulfilling physicians’ obligations to protect the well-being and privacy of their patients is complicated in the age of the internet. Internet culture is very different from that of the medical profession, creating potential ethical problems with appropriate boundaries and privacy that did not exist when physicians interacted exclusively offline. In order to maintain the trust of the public and that of individual patients, physicians increasingly need to understand the limits and risks of disclosure of certain types of information online. Although concerns about unprofessional medical student and resident behavior online have been articulated before,^5,6^ the ethical risks of public disclosure, when narrative medicine intersects with social media, remain poorly defined.^7,8^

In May 2018, thousands of individuals—including many health care professionals— shared health-related stories on Twitter using the hashtag #ShareAStoryInOneTweet in response to one physician’s spontaneous tweet about a patient story, with this hashtag included. Certain tweets included potentially identifiable information that could be considered a breach of confidence if disclosed without patient consent, risking harm to patients, physician’s careers, and public trust in the profession.

A July 2018 article highlighted the importance of sharing stories but did not address the potential risks of sharing online.^9^ Over time, some ‘viral’ tweets were deleted, raising further concerns that the easy platform for disclosure might have led to posts that authors subsequently regretted. A notable example of a popular post (altered to avoid identification) was retweeted 13,491 times and liked by 55,994 people before being deleted:

> *“I delivered a baby very underweight, weighing two pounds. They said he did not have a chance. I remained with him for a couple of days. Nine years later, he played his first football game last week*.”

Hashtags can make online content searchable and discoverable online, regardless of time since publication. The American Medical Association, Massachusetts Medical Society, and other organizations advise physicians to report unprofessional social media use.^10,11^ What constitutes unprofessional behavior on social media is not clearly defined. To advance a common understanding and to facilitate subsequent discussion within the profession about what is appropriate, we sought to describe participation of physicians and other health professionals in this event, the reach of their postings, and the prevalence of potentially identifiable disclosures about patients.

## Methods

### Cohort Definition

Because all information about the published content were publicly accessible, Lowell General Hospital approved this study as IRB exempt. To evaluate content in the #ShareAStoryInOneTweet phenomenon, we used Symplur Signals, a proprietary healthcare-focused database and analytics program collecting data on Twitter using its Enterprise application program interface.^12^ The first tweet with the hashtag occurred May 4, 2018. From May 1, 2018 through December 31, 2018, we identified 45,040 tweets including the hashtag #ShareAStoryInOneTweet. We focused on the 43,374 tweets shared in the month of May 1 2018 00:00:00 to June 1 00:00:00 Eastern Standard Time. We conducted our analysis in January 2019, eight months after the study period.

We used the software program’s classification to identify doctors, patients, and other healthcare stakeholders (e.g. caregivers, pharmaceutical firms, academic or research organizations) based upon public self-identification in their Twitter profiles.^13^ For May 2018, we identified 4,871 tweets with the hashtag shared by doctors and other healthcare providers, of which 1,206 were unique (the remainder represented “retweeting” of prior postings).

We reviewed the 1,206 tweets by reading text provided within the dataset and then evaluating the URL and each account’s public profile on Twitter’s website as of March 2019. We excluded those from students misclassified as healthcare providers (1.3%) (e.g. listing ‘future doctor’ in profile), blocked accounts (0.3%), or if no relevant content was posted with the hashtag (1.2%), leaving a total of 1,172 tweets shared by physicians, nurses, or other health professionals [**Figure 1**]. We excluded 26 “retweets” in which the authors used the hashtag to share someone else’s disclosure rather than their own and 127 that included the author’s own illness experiences rather than those of others. We also excluded 78 tweets (6.5%) with content found in the dataset but deleted from Twitter when evaluated on the website.

**Figure 1.**
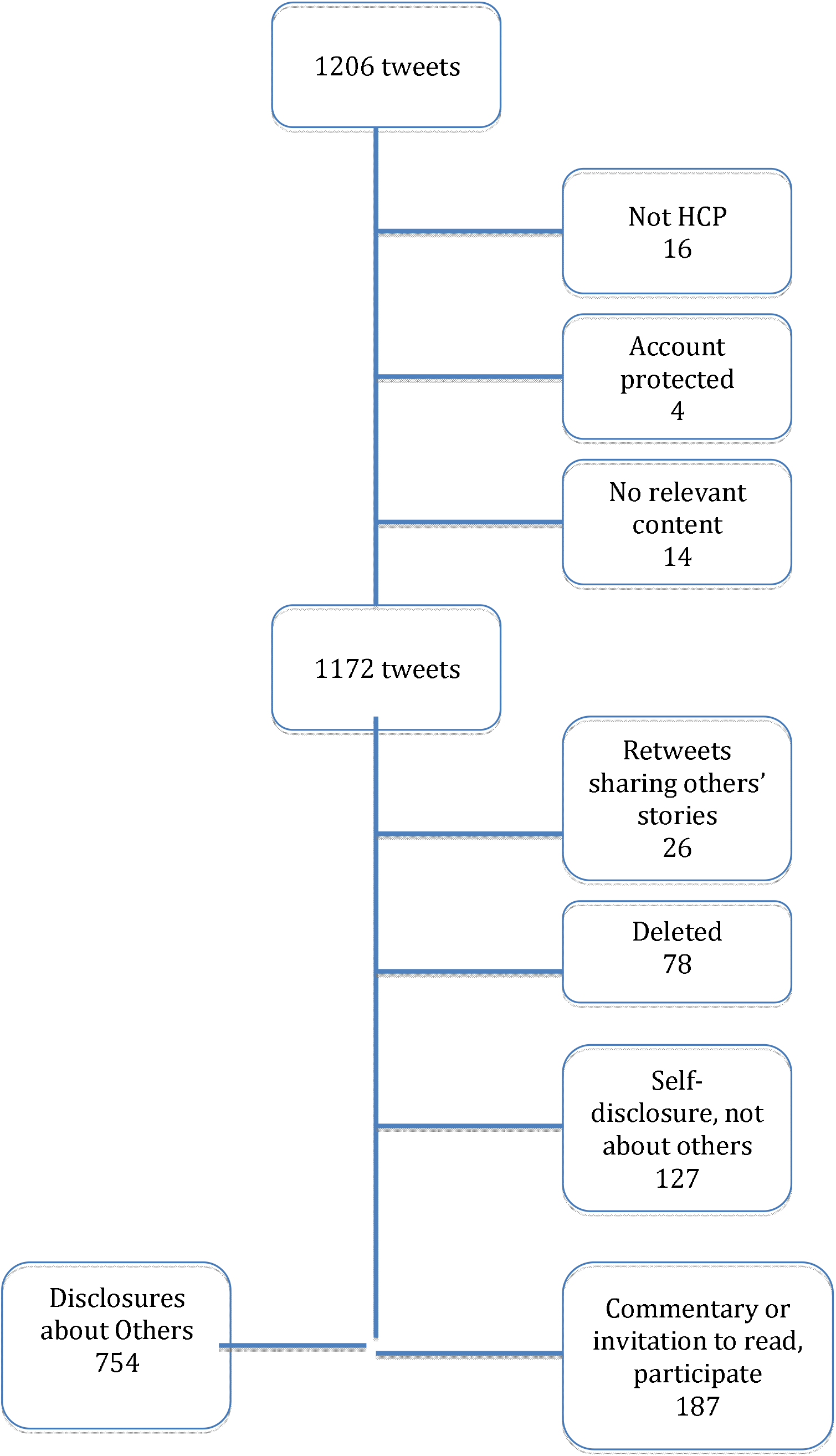
Tweets by doctors and other health care providers (HCP) during #ShareAStoryInOneTweet in May 2018.

We examined characteristics of the 656 health professionals sharing these tweets, using information publicly available in their online profiles, including profession, gender, and country. For physicians, we also categorized by specialty as described in their profiles, or unknown if not stated. More detailed content analysis focused upon the 754 tweets (62.5%) in which the healthcare professional shared the illness or clinical experience of someone other than the tweet author.

We also evaluated the 187 tweets commenting upon the hashtag-related phenomenon or recommending participation to others separately. We analyzed tweets individually rather than as content threads.

### Measures

To assess the magnitude of hashtag use, we evaluated total number of tweets. We also evaluated the number of total participants, focusing upon physician, nurse, and other HCP use during May 2018. We calculated tweets per hour, number of tweets, and use of images both in aggregate and by healthcare stakeholder categories. In order to capture hourly tweet activity rates, we restricted the time frame to the first two weeks starting May 4^th^ to focus on the ‘viral’ period of sharing. We evaluated the potential reach of the tweets using the software’s definition of impressions, calculating each tweet author’s follower count at the time of each tweet’s publication online (e.g. a doctor posting while having 500 followers = 500 impressions).

For the content analysis, for each tweet we coded several measures: the tweet author’s role in the other person’s clinical care; whether the patient died or was actively dying; whether the author helped save the other person’s life; inclusion of patient name; inclusion of a clinical image; and inclusion of a specific age. Based upon the tweet, we categorized the time frame of the event(s) described as within the past year, 1-2 years ago, 2-5 years ago, >5 years, or unknown.

To assess whether a either a patient or her/his family or friends would be able to identify the clinical scenario described in each tweet, we categorized both questions broadly as likely (defined as “more likely than not to be identifiable”) or not (not meeting that threshold). One author (MK) assessed all tweets; a second author (WA) coded a 10% sample independently and inter-coder reliability was assessed with ReCal software.^14^ The two authors then reached consensus on discrepancies and used this exercise to identify any areas where the first coder might systematically have erred.

Because the tweets could be discoverable in malpractice or tort suits, we also analyzed whether the author made comments with a negative opinion about the patient or family, or if s/he acknowledged that a medical error occurred. We also assessed whether information was shared about vulnerable patients, as defined by the U.S. Department of Health and Human Services.^15^

We separately evaluated the 187 tweets commenting upon the hashtag to determine whether the authors had a favorable or unfavorable opinion of the viral sharing, or if they invited others to share stories or to participate. We also identified whether these tweets expressed any concern about privacy breaches.

### Statistical Analysis

Symplur provided data for overall activity and frequencies for stakeholder participation. For the detailed content analysis we calculated frequencies, median and mean endpoints with Microsoft Excel for Mac 2011 version 14.7.2. We used Cohen’s kappa statistic to measure inter-rater reliability.^16^

## Results

### Tweet Volume and Participants

For May 2018, we identified 31,690 participants posting tweets with the hashtag, with a potential of 106.5 million views. A total of 1725 individuals self-identifying as doctors and 861 as other healthcare providers shared tweets with the hashtag, representing 5.3% and 2.6% of the total participants, respectively. At its peak activity for sharing, participants in this phenomenon tweeted 1274 times/hour **[Figure 2]**.

**Figure 2.**
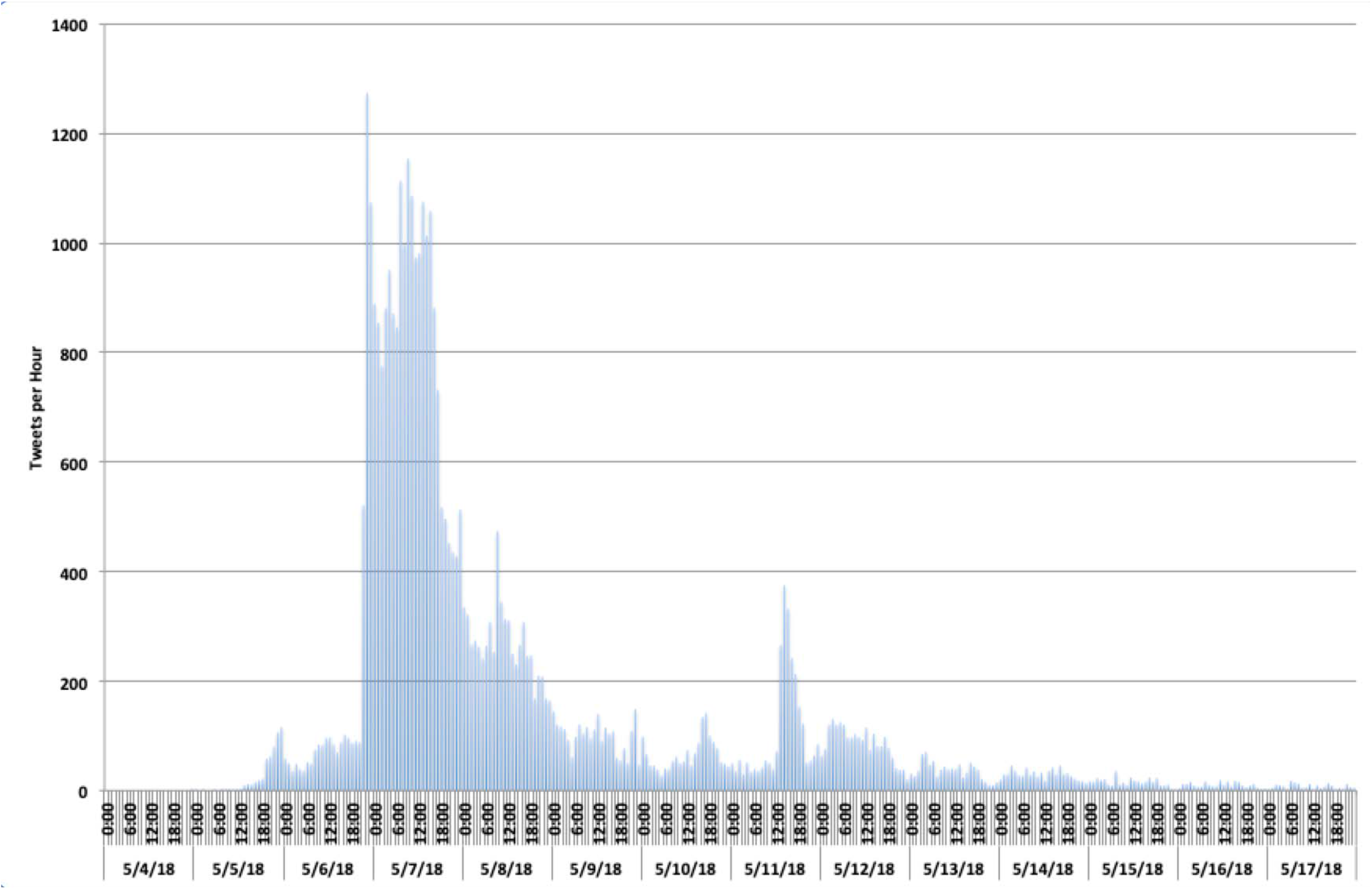
Tweets per hour including the hashtag #ShareAStoryInOneTweet, May 2018.

### Participants and Tweets with Disclosures about Others

The characteristics of the 656 health professionals sharing tweets with disclosures about others are presented in **Table 1**. Of the participants, 384 (58.5%) were female. Physicians constituted the largest proportion of the tweeters (74.1%), with nurses representing a minority (14.9%). Among physicians, emergency medicine, family medicine/general practice, and hematology-oncology were the specialties most frequently represented. Most of the 656 participants were in the United States (52.9%), followed by Canada (15.1%) and the United Kingdom (12.5%).

**Table 1.** Characteristics of 656 doctors and healthcare providers sharing 754 tweets disclosing information about others

| **Participants** |  | **No.** | **%** | **% of subgroup** |
| --- | --- | --- | --- | --- |
| *Gender* |  |  |  |  |
|  | Female | 384 | 58.5% |  |
|  | Male | 266 | 40.5% |  |
|  | Unknown | 6 | 0.9% |  |
| *Profession* |  |  |  |  |
|  | Doctor | 486 | 74.1% |  |
|  | Anesthesia | 22 |  | 4.5% |
|  | Cardiology | 25 |  | 5.1% |
|  | Critical Care | 12 |  | 2.5% |
|  | Emergency Medicine | 77 |  | 15.8% |
|  | Family Medicine-General Practitioner | 48 |  | 9.9% |
|  | Gastroenterology | 6 |  | 1.2% |
|  | Hematology-Oncology | 39 |  | 8.0% |
|  | Hospitalist | 5 |  | 1.0% |
|  | Infectious Disease | 5 |  | 1.0% |
|  | Internal Medicine | 12 |  | 2.5% |
|  | Neurosurgery | 5 |  | 1.0% |
|  | OB/GYN | 13 |  | 2.7% |
|  | Palliative Care | 18 |  | 3.7% |
|  | Pathology | 9 |  | 1.9% |
|  | Pediatrics | 28 |  | 5.8% |
|  | Psychiatry | 7 |  | 1.4% |
|  | Pulmonary Medicine | 7 |  | 1.4% |
|  | Radiation<br>Oncology/Clinical<br>Oncology | 10 |  | 2.1% |
|  | Radiology | 6 |  | 1.2% |
|  | Surgery | 19 |  | 3.9% |
|  | Trauma Surgery | 20 |  | 4.1% |
|  | Unknown | 36 |  | 7.4% |
|  | Other | 57 |  | 11.7% |
|  | Nurse | 98 | 14.9% |  |
|  | Nurse, NOS | 66 |  | 67.3% |
|  | Critical Care | 16 |  | 16.3% |
|  | Emergency Medicine | 8 |  | 8.2% |
|  | Other | 8 |  | 8.2% |
|  | Nurse Practitioner | 12 | 1.8% |  |
|  | Paramedic | 18 | 2.7% |  |
|  | Pharmacist | 7 | 1.1% |  |
|  | Physical Therapy | 10 | 1.5% |  |
|  | Psychologist | 5 | 0.8% |  |
|  | Social Worker | 7 | 1.1% |  |
|  | Speech Therapy | 5 | 0.8% |  |
|  | Other | 8 | 1.2% |  |
| *Country* |  |  |  |  |
|  | Australia | 11 | 1.7% |  |
|  | Canada | 99 | 15.1% |  |
|  | India | 7 | 1.1% |  |
|  | Ireland | 16 | 2.4% |  |
|  | Saudi Arabia | 7 | 1.1% |  |
|  | South Africa | 4 | 0.6% |  |
|  | Spain | 4 | 0.6% |  |
|  | United Kingdom | 82 | 12.5% |  |
|  | United States | 347 | 52.9% |  |
|  | Unknown | 55 | 8.3% |  |
|  | Other | 24 | 3.7% |  |

Among the 754 tweets focused upon the illness experience of someone other than the author, the majority (87.4%) involved the sharing of stories about direct patient care, rather than the author’s role as a family caregiver or in another role **[Table 2]**. In 13.6% of cases, tweets included a specific time frame. The patient’s age was included in 163 (21.6%) of these tweets, and patient name was included in 15 (2.0%). Eleven tweets (1.5%) shared a clinical image. 152 tweets (20.2%) shared information about people in a vulnerable category. Two posts (0.3%) mentioned patient consent to share within that tweet, one explicitly and one inferred from past patient agreement to share a specific story. Based upon the number of likes, a minimum of 154,900 accounts viewed these 754 tweets.

**Table 2.** Content Characteristics of 754 Tweets with Disclosures about Others

|  |  | No. (%) | % |
| --- | --- | --- | --- |
| Author Role |  |  |  |
|  | Healthcare Professional | 669 | 88.7% |
|  | Patient | 0 |  |
|  | Caregiver | 42 |  |
|  | Other | 43 |  |
| Time Frame Described |  |  |  |
|  | Within past year | 5 | 0.7% |
|  | 1-2 years | 6 | 0.8% |
|  | 2-5 years | 5 | 0.7% |
|  | > 5 years | 79 | 10.5% |
|  | Unknown/Not Described | 659 | 87.4% |
| Content |  |  |  |
|  | Author involved in patient care | 635 | 84.2% |
|  | Actively dying patient or patient death | 337 | 44.7% |
|  | Saving a patient's life | 131 | 17.4% |
|  | Include a clinical image | 11 | 1.5% |
|  | Include patient name | 15 | 2.0% |
|  | Provide specific patient age | 163 | 21.6% |
|  | Express negative opinion of patient or family | 4 | 0.5% |
|  | Mention medical error | 6 | 0.8% |
| Estimated Likely ("more likely than not") |  |  |  |
|  | Can family or friends identify situation described? | 242 | 32.1% |
|  | Can patient identify situation described? |  |  |
|  | *All tweets* | 183 | 24.3% |
|  | *Potentially living patients (348 tweets)* | 162 | 46.6% |
| Vulnerable population |  | 152 | 20.2% |

Nearly half of the tweets (44.7%) described a clinical scenario involving death or dying. Comments disclosing medical errors or expressing a negative opinion about the patient or family were rare (0.8% and 0.5%, respectively).

Inter-coder reliability in rating the identifiability by patients or by family both demonstrated 86.8% agreement, with a Cohen’s Kappa of 0.8 suggesting substantial agreement. In evaluating the potential for unintended identification of the clinical scenario, we estimated that almost one third (32.1%) of families or friends would likely find the content in the tweet identifiable. Among patients who were potentially still living, we estimated that nearly half (46.6%) likely could identify the information shared about themselves. Of the 162 tweets we estimated patients would likely be able to identify, we estimated that 81 (50%) would likely be identifiable by families and friends. The 754 tweets by participants received a median of two retweets (range: 0-19,959) and 16 likes (range: 0-56,735).

### Tweets Relating to the Hashtag

Of the 187 tweets actively part of the conversations without disclosures, 173 (92.5%) made some commentary. Six (3.2%) raised concerns about privacy or identifiable information in the tweets with disclosures, and one involved another critical comment; the remaining 167 were neutral or favorable. A total of 42 tweets (22.5%) invited others to read the hashtag’s stream or contribute to it.

## Discussion

In this retrospective study, we describe a physician-initiated event sharing health-related stories and information on Twitter, quantifying the global participation of healthcare professionals and the type of content shared. Their tweeted stories became widely shared, attracting media attention and disseminating the information widely. Almost none explicitly appear to confirm consent to share information publicly on a popular social network. Nurses, physicians, and other professionals commenting using the hashtag were more likely to express support for the event and encourage others to participate than they were to raise concerns about patient privacy breaches. However, recent research suggests that 12% of patients may have less trust in physicians describing patient stories on social media, even if shared respectfully.^17^

We observed a relatively high incidence of sharing stories including details that might make them potentially identifiable to patients themselves and even to families and friends in a setting that involved a large number of healthcare professionals compared to prior studies. Early in the use of social media, most U.S. state medical boards received at least one episode of online professionalism violations reported for disciplinary action, including violations of patient confidentiality.^18^ Surveys of medical students and physicians suggest the prevalence of unprofessional behavior among medical students is infrequent.^19,20^ Our study indicates that in some circumstances healthcare professionals may share more information publicly than the public might expect. Privacy breaches risk potential negative effects for relationships with their patients, for professional disciplinary actions or torts, and for erosion of public trust.

Our findings differ from prior studies of online medical professionalism at least partly because we analyzed in detail a specific event focused upon health-related disclosures. There is no indication this episode was planned, but the incidence of similar episodes is unknown. But it is not an isolated event; for example, another prominent example involves physicians opposing gun violence, who used the hashtag #ThisIsOurLane on Twitter in November 2018.^21^ Physicians focused on policy issues, but some may have failed to recognize privacy concerns, publishing tweets with photographs similar in nature to prior social media content that has in certain other cases resulted in professional termination.^22,23^ Social media studies publishing tweets often permit reverse identification of the authors,^24^ and a survey suggests that most ‘participants’ are somewhat or very uncomfortable if their tweets were to be quoted in a published research paper.^25^ In our study, we found that 6.5% of tweets archived in the software’s archived dataset were in fact deleted by healthcare professionals, indicating that some did not want their tweets to remain publicly visible. Even if deleted online, publishing tweets in journals enhances permanence and discoverability.

Most research evaluating online disclosures focus on the privacy paradox, in which people value their privacy but still share their own information. Surveys indicate people may value short-term social rewards of self-disclosure online more than long-term privacy concerns,^26^ and high social capital of social network users is associated with increased self-disclosures over time.^27^ For people disclosing information about others, the research is more limited, but opinion leadership and female gender are linked to less concern about others’ privacy,^28^ consistent with the findings of our study. Healthcare professionals may be prone to these same tendencies, despite their training and education to maintain privacy. Generational differences concerns about privacy online may also play a role,^29^ but we could not assess that possibility in this study.

Based upon the temporal pattern of sharing, this hashtag-related event may be less similar to narrative medicine and writing than to a brief episode of social contagion, in which viral sharing of content or emotions online may occur and involve more than simple, conscious risk/reward tradeoffs.^30,31^ Unlike traditional peer-reviewed publication of a medical story in narrative medicine, tweeting occurs quickly and does not permit editing. The observation that 6.5% chose later to delete their contributions may suggest that some health care professionals who participated in the experience may have later viewed their behavior as a temporary lapse in judgment.

Another contributing factor may be a knowledge gap for physicians and other healthcare professionals on how to behave online. While many recognize the importance of online professionalism, curricula for formal medical education training are only beginning to emerge and remain uncommon.^32-4^ In this study, no tweet with met criteria for justifiable release of identifiable information without consent, and the ethical obligation to maintain confidentiality does not end with a patient’s death.^35^ The digital medium does not avoid the potential that disclosures about patients risk breaching confidentiality, undermining trust within that therapeutic relationship as well as public trust in the medical profession. Our findings suggest a potential need for evidence-based training in ethical digital communications skills for the undergraduate, graduate and continuing medical education. Professional societies could create resources that allow social media authors to document having obtained consent, so that disclosing identifiable patient information without consent does not inadvertently become normalized.

### Limitations

Our study has several limitations. First, we studied a very specific event that may occur during very active periods of online engagement but likely overestimates the general incidence of online behaviors that could in some cases constitute violations of medical professionalism. Second, we could not assess the number of people actually seeing these tweets, resulting in a wide range since we could only measure the number of likes and potential reach. Third, we only analyzed tweets from accounts that the software identified as healthcare professionals. Our evaluation of all tweets in the cohort confirms the software rarely misclassified non-professionals into this group, but we did not evaluate any other participants in the event to determine if we could identify more participating healthcare professionals not categorized as doctors or nurses by the software, which could decrease or increase the incidence of potential privacy breaches. Fourth, by analyzing only tweets with the hashtag, we potentially underestimated the frequency of others expressing concern about patient privacy. Fifth, given the brevity inherent to the medium of Twitter, it is possible that some authors did indeed have formal documentation of patients’ consent to share their stories but that there was insufficient room to include due to character limits in each post. Finally, our assessment of identifiability might differ from others, and we cannot exclude the possibility that some physicians and nurses tweeting what seemingly identifiable stories consciously changed important details to de-identify. It was beyond the scope of this study to confirm whether any harm occurred.

Despite these limitations, our study clearly shows that internet-based sharing raises potential pitfalls for medical professionalism. The Internet provides nurses, physicians, and other professionals the opportunity to help or harm others on a global scale. Although Internet culture may favor maximizing transparency, it can also pose the risk of directly contradicting health professionals’ fiduciary duty: first, do no harm, including in what we say.

## Conclusion

We identified an episode with a high incidence of potential privacy breaches online compared to previous reports. More research is essential to confirm our findings and determine how to ensure physicians, nurses, and other professionals adapt their behavior to maintain medical professionalism in the digital age.

## Data Availability

All data were obtained from the Twitter API through analytics software, Symplur Signals.

## Author Contributions

Dr. Katz had full access to all of the data in the study and takes responsibility for the integrity of the data and the accuracy of the data analysis. All estimates and analyses in this article are by the authors.

Concept and design: Ahmed, Gutheil, Jagsi, Katz

Acquisition, analysis, or interpretation of data: Ahmed, Gutheil, Jagsi, Katz

Drafting of the manuscript: Katz, Jagsi

Critical revision of the manuscript: Ahmed, Gutheil, Jagsi, Katz Statistical analysis: Katz

Obtained funding: Katz

Administrative, technical, or material support: Katz

Supervision: Gutheil, Jagsi

